# Factors associated with the sero-prevalence of Rickettsioses in Northern Tamil Nadu, India

**DOI:** 10.64898/2026.03.24.26348878

**Authors:** Solomon D’Cruz, Sreedevi Kottamreddy, Teena Mariam Thomas, Karthik Gunasekaran, Susmitha Karunasree, John Antony Jude Prakash

## Abstract

**Introduction:** Rickettsial infections are vector-borne diseases and sero-prevalence shows regional and population-based variation. This prospective study assesses the sero-prevalence and factors associated with rickettsial infections in different geographical regions of Northern Tamil Nadu, India.

**Methods:** A community-based cross-sectional study was performed among 2565 individuals in five districts in Tamil Nadu who consented and provided 4 ml blood samples. A semi-structured questionnaire was administered to gather information on socio-demographic and environmental parameters. The serum IgG antibodies to scrub typhus (ST), spotted fever (SF), murine typhus (MT) and Q fever (QF) were detected by ELISA and the optical density (OD) recorded. Epi-Data was used for data entry and SPSS for analysis.

**Results:** The sero-prevalence of ST, SF, MT and QF was 14%, 9.1%, 3.7%, and 5.7%, among the study population. For ST, age >35, people living in rural and <1000 MSL, farmers, sleeping on the floor, lack of toilet at home, and grass near the home were associated with higher risk. For SF, people living at 501 – 1000 MSL elevation, no toilet at home and having pet animal were associated with higher odds. Females, 46 – 55 and >65 years, people residing in urban areas and elevation upto 500 MSL, those who sleep on the floor, not changing clothes daily and with pet at home were associated with higher odds of MT. Whereas Q fever was more likely to affect who sleep directly on the floor, residing in urban and do not have pets.

**Conclusion:** Scrub typhus is the most common rickettsial infection followed by spotted fever in Northern Tamil Nadu. Factors associated with prevalence vary for different rickettsial diseases and include personal and lifestyle behaviors. The findings of this study need to be verified by multi-centre cohort studies.

## Introduction

Rickettsial illnesses, including Scrub Typhus (ST), Spotted Fever (SF), and Murine Typhus (MT), are transmitted via vectors (1), whereas Q fever (QF) is predominantly transmitted by aerosols and less often by vectors (1). The agents causing rickettsial infections belong to the genera *Rickettsia, Orientia* and *Coxiella* (2). ST is caused by *Orientia tsutsugamushi*, SF by SFGR (spotted fever group rickettsia), and MT by *Rickettsia typhi* (3,4). The trombiculid mite (Chigger mite) is the vector responsible for the spread of ST, while ticks are the causative agents of SF and fleas transmit MT. Q fever (QF) is a caused by the bacterium *Coxiella burnetii* and transmitted mainly by inhaling aerosols from infected animal fluids and less commonly by ticks (3). Rickettsial infections are major threat to public health as there is a low degree of clinical suspicion which is compounded by the lack of diagnostic facilities and capabilities (5). In South East Asia, rickettsial infections are the second commonest cause of acute undifferentiated febrile illness (AUFI) next to dengue (6) and it is the most common AUFI in Vellore (7).

Among all the rickettsial infections, scrub typhus is the commonest in India accounting for a sero-prevalence of 34.2% among the community (8). The sero-survey done in East India (Odisha) has reported a sero-prevalence of 56.2% of scrub typhus (9), among the healthy blood donors in Vellore it was 15% (10). Despite its high sero-prevalence, scrub typhus is often underdiagnosed as it presents like any other acute undifferentiated febrile illness especially when the eschar is not looked for or is absent (11). Other rickettsial infections which have been reported from India are spotted fever and murine typhus which present as AUFI with rash (12). Q fever primarily affects individuals employed in the livestock sector, involving cattle, sheep, and goats and often presents as self-limiting febrile illness (13).

Studies from different regions have identified various risk factors for scrub typhus. In Vellore, farming or gardening, presence of water bodies (100m), outdoor cooking, owning a pet, bushes within 5m, and not drying clothes on a clothesline were associated with higher risk for scrub typhus (14,15). A study from Odisha reported that presence of swampy areas, firewood piles near houses, construction work, and frequent visits to bushy and lying or sitting on grass increased the risk for scrub typhus (16). A retrospective analysis further showed that females, individuals over 50 years, and those working as farmers, daily wage laborers, or housewives were more likely to be affected (17). The preliminary analysis of the same data has revealed that increase in age, people with primary education, agricultural workers were at increased risk for scrub typhus (18).

A sero-prevalence study from Bhutan reported that people aged >40 years had a higher odds of spotted fever positivity (3). Another study among pastoralist community in Northern Tanzania in 2024 revealed that herding cattle and increase in age were positively associated with the of spotted fever (19). A study conducted in Vientiane, Lao PDR, reported that individuals who had spent more than one-third of their lifetime in the area, retail traders, unemployed persons, those living close to markets, and residents of neighborhoods with high building density were at higher risk of past murine typhus infection (20). Agricultural workers and people living close to the forest areas were at high risk for spotted fever sero-positive in the study published with the preliminary data (18). This study was conducted to determine the sero-prevalence and the factors associated with the sero-positivity of rickettsial infections in five districts of Tamil Nadu, South India.

## Methodology

### Study type and Study setting

During the community based cross-sectional sero-survey (previously published) undertaken in Erode, the Nilgiris, Salem, Tiruvannamalai and Vellore districts of Tamil Nadu (21), a detailed questionnaire was administered. The study was performed from September 2017 to January 2020. This study includes participants living in plains, highlands, people from urban, rural and peri forested areas and those with diverse occupations. Agriculture is the major occupation in all the 5 districts surveyed with varying crops being cultivated across the districts. Even though the districts surveyed are adjacent and share boundaries with each other, they vary drastically in terms of irrigation facilities, crops, climate, forest cover, etc.

The sample size was calculated using OpenEpi software, the margin of error was 5% and the confidence interval was set at 95%. A design effect of 3 was considered and the final sample size required for the study was 2,399. The detailed methodology of this study, including participant selection, eligibility criteria, and sampling techniques, has been described in a previously published article (21). After obtaining informed consent, a questionnaire was administered to the participants, the questionnaire included demographic details and risk factors for acquiring rickettsial infections. In addition to the demographic data, information regarding the personal habits, vegetation around the house and pet animals were collected. Elevation profile in meters above sea level (MSL) of each locality was obtained from Bharat Maps (https://bharatmaps.gov.in/newversion/map.aspx). The shape files were downloaded from the Survey of India (https://surveyofindia.gov.in/) and Q GIS software 3.38.3 was used for spatial analysis.

### Detection of IgG antibodies to Scrub Typhus, Spotted fever, Murine Typhus and Q fever

From enrolled participants, 4 ml blood was collected in BD Vacutainer Clot Activator Tube (BD, Plymouth, UK). The serum was separated by centrifugation and tested for detection of IgG antibodies to Scrub Typhus, Spotted fever, Murine Typhus and Q fever at a dilution of 1:100. IgG antibodies to O. tsutsugamushi were detected using the Scrub Typhus Detect IgG ELISA system (InBios International Inc., Seattle, WA). While IgG antibodies to murine typhus and spotted fever were detected by R. typhi IgG ELISA (Fuller Laboratory, Fullerton, CA) and the Rickettsia conorii ELISA IgG/IgM (Vircell, Granada,Spain). As described previously, serum sample was considered to be positive if the OD was ≥1.5 for any of these infections (18). Q Fever Phase 2 IgG antibodies were detected using the NovaLisa *Coxiella burnetii* (Q-fever) Phase 2 IgG ELISA (NovaTec Immundiagnostica GmbH, Dietzenbach, Germany). All the assays were performed with appropriate controls on an automated ELISA workstation (Euroimmun Analyzer I, Euroimmun AG, Lubeck, Germany). The OD cut-off for Q Fever was set at 0.6 and above since the mean plus 2 SD was just above 0.6 (Data not shown).

### Statistical methods

Data entry was performed using EpiData 3.1 (Jens M. Lauritsen, Odense, Denmark), and statistical analysis was carried out using IBM SPSS Statistics for Windows, Version 21.0 (IBM Corp., Armonk, NY, USA). The chi-square test was used to assess the association between categorical variables, and unadjusted odds ratios (ORs) with 95% confidence intervals (CIs) were reported to measure the strength of association. Logistic regression analysis was performed to adjust for potential confounders, and adjusted odds ratios (AORs) with 95% CIs were presented. A p-value of <0.05 was considered statistically significant.

### Ethics Approval

The study was approved by the Institutional Review Board (IRB) and Ethics committee of Christian Medical College, Vellore (IRB Min. No. 9369 dated 25th March 2015).

## Results

In this prospective study, 2565 samples were studied from the five Northern Tamil Nadu districts surveyed (Refer Figure 1). Majority of the samples were collected from females (60%) and the age group of 31 – 45 years (31.5%). Daily wage workers contribute to half of the study population with middle and high school education being the educational level of the majority.

**Figure 1:**
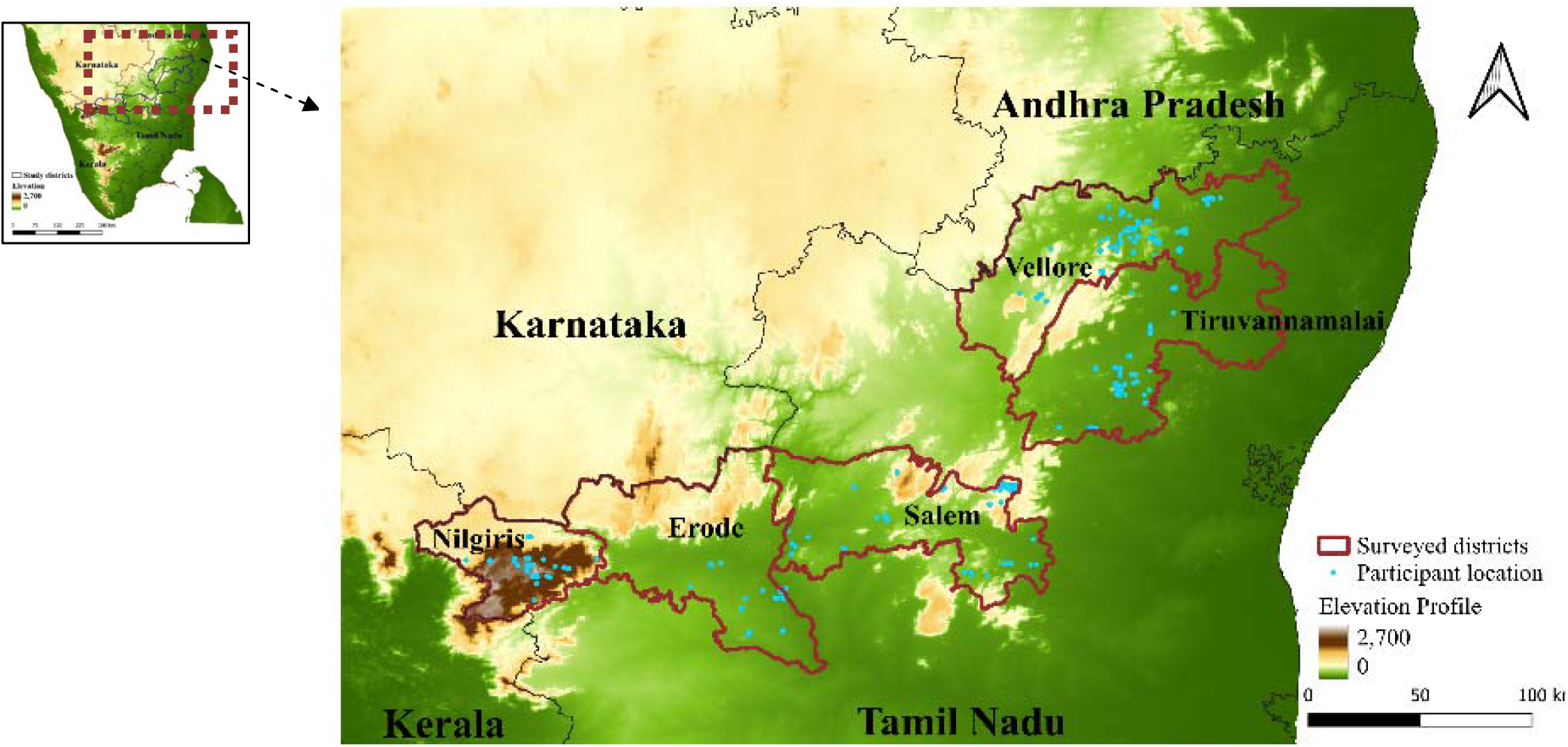
Elevation profile: Geographic location of the study participants.

Fifty percent of the study population is from rural areas and most of the population are from areas of less than 500 meters in elevation (Refer Supplementary Table 2 for details). The elevation profile of the study areas with the geo-coded location of all study participants are shown in Figure 1.

### Sero-prevalence of rickettsial infections across the study districts

The sero-prevalence of ST in our study districts ranges from 2% (The Nilgiris) to 22.1% (Tiruvannamalai), SF ranges from 0% (Erode) to 19.9% (Salem), MT ranges from 1.7% (Vellore) to 10.5% (Tiruvannamalai) and QF ranges from 1.3% (Tiruvannamalai) to 11.5% (Salem). The actual seroprevalence of all four rickettsial infections across the surveyed districts is as given in Supplementary Table 1 and it is depicted in Figure 2. The geo-coded location of the positives (ST, SF, MT & QF) are shown in the Supplementary Figure 1.

**Figure 2:**
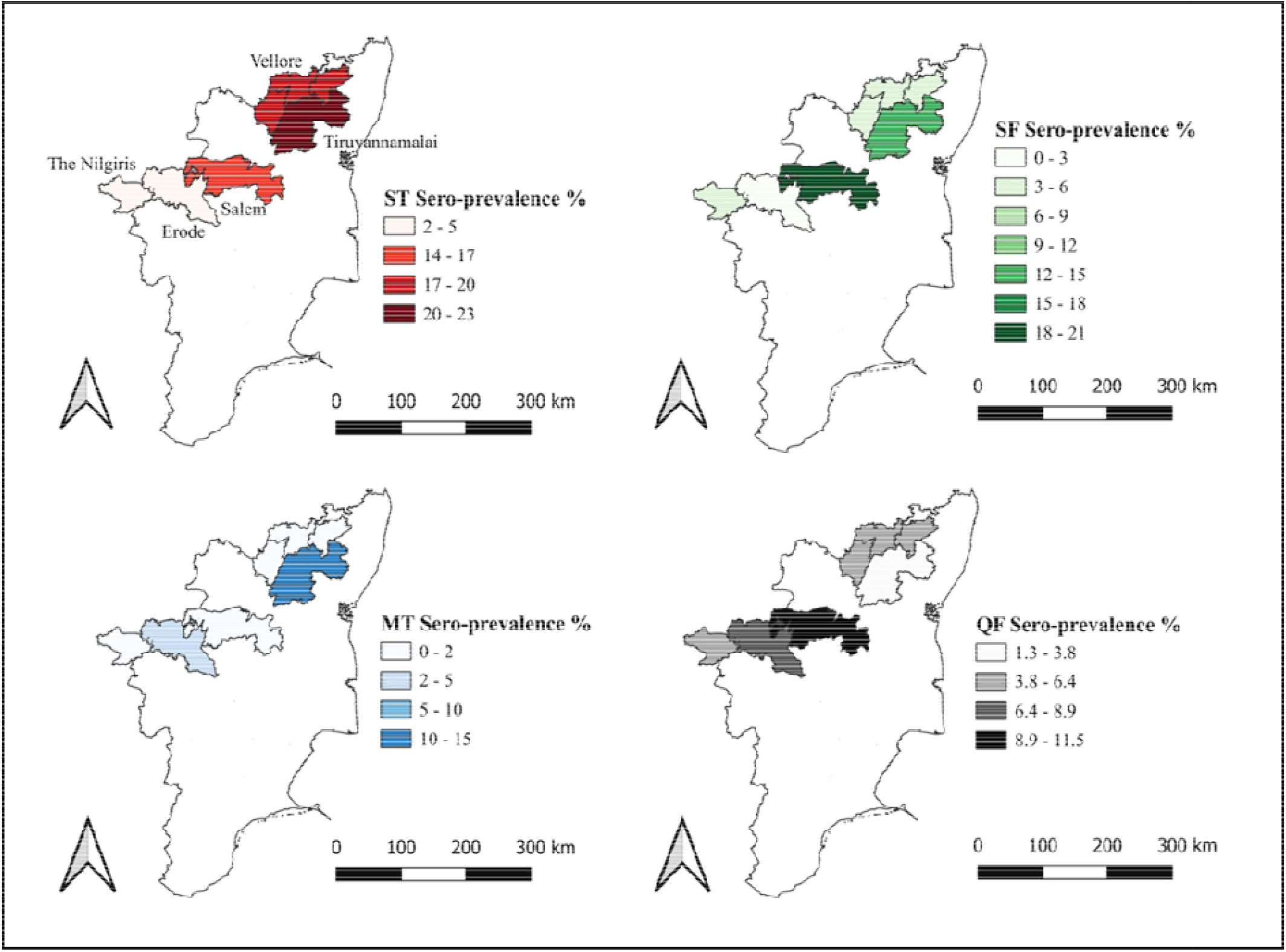
Sero-prevalence of rickettsial infections in the surveyed districts.

### Univariate analysis

The detailed univariate analysis of the risk factors and the rickettsial infections are given in the supplementary tables (Supplementary Table 3 to 6). When compared to males, females had the higher odds of being sero-positive for ST and MT, there was no gender association with SF and QF. Age was strongly associated with the scrub typhus positivity, the risk increased with the increase of age. Whereas for SF, only the age group of 46 – 65 years were at risk, for MT, it is 46 – 55 years and for Q fever it is >65 years when compared to 16 – 25 years. With people living in urban as reference, people from rural areas were at higher odds of being positive for ST and people from rural and peri-forested areas were at higher odds of being positive for SF. On the other hand, people from urban had higher association with MT and QF positivity when compared with people from peri-forested areas. With more than 8 years of education as reference, people with 8 years or less of education are at higher risk for ST and SF. People with 0 years of education were at high risk for QF. People living in elevation of 0 – 1000 meters above sea level (MSL) seems to be highly associated with ST positivity and 501 – 1000 MSL for SF when compared to >1000m above sea level. Occupation like farming, daily wages and housewives were highly associated with ST and only daily wage workers for SF when compared with other occupations. Other statistically significant risk factors for ST are people sleeping on ground with or without mat/mattress, not having toilet inside the house, owning a pet animal at home and having grass and bushes near house. For SF, the significantly associated risk factors are people sleeping on ground with or without mat/mattress, not having toilet at home, not changing clothes daily after work, owning a pet animal and has grass and bushes near house. Whereas for MT, it is people sleeping on ground with mat/mattress and owning a pet animal. On the contrary, for QF it is people who do not own a pet animal and people sleeping on ground are at risk.

### Multivariate analysis

When subjected to multivariate analysis and adjusted for confounders, for ST, people who were aged above 35 years were at higher odds of being positive when compared to 16 – 25 years of age. People aged 46 to 55 and >65 years were at higher risk for MT. SF, and QF, on the other hand, were not significantly related to age. People from rural areas were at significant risk for ST when compared with people from urban areas. On contrary, for MT and QF, when compared to peri-forested areas, people from urban had higher risk to be sero-positive. Elevation was proved to be significant risk factor for ST, SF and MT as the people living at an elevation of 0 – 1000 MSL had higher odds for ST positive, 501 – 1000 MSL for SF and 0 – 500 MSL for MT and when compared to people living at an altitude of 1000 MSL. Farmers was a significant risk factor only for ST when compared to other occupations. Sleeping on ground with or without mat/mattress was significantly related to ST positivity, whereas for MT it is sleeping on ground with mat/mattress and for QF it is sleeping on ground without mat/mattress. Not having toilet at home was significantly related to ST and SF positivity. Not changing house dress daily was a risk factor for MT sero-positivity. Having pet animal at home was significantly related with SF and MT positivity; however, not having a pet at home was significantly associated with QF positivity. Having grass land near house was a risk factor for ST positivity.

**Table 2:** Factors associated with the prevalence of ST, SF, MT & QF.

| Variables<br>(n = 2565) | Scrub Typhus |  | Spotted Fever |  | Murine Typhus |  | Q Fever |  |
| --- | --- | --- | --- | --- | --- | --- | --- | --- |
|  | AOR<br>(95% CI) | p<br>Value | AOR<br>(95% CI) | p<br>Value | AOR<br>(95% CI) | p<br>Value | AOR<br>(95% CI) | p<br>Value |
| **Gender** |  |  |  |  |  |  |  |  |
| Male | **Reference** |  |  |  |  |  |  |  |
| Female | 1.10<br>(0.83, 1.46) | 0.512 | 0.81<br>(0.58, 1.11) | 0.186 | **2.38**<br>**(1.37, 4.13)** | **0.002** | 0.70<br>(0.47, 1.04) | 0.081 |
| Age group in years |  |  |  |  |  |  |  |  |
| 16 – 25 | Reference |  |  |  |  |  |  |  |
| 26 – 35 | 0.84<br>(0.50, 1.43) | 0.524 | 1.37<br>(0.76, 2.47) | 0.290 | 1.97<br>(0.82, 4.76) | 0.131 | 0.91<br>(0.44, 1.88) | 0.800 |
| 36 – 45 | **1.70**<br>**(1.02, 2.81)** | **0.040** | 1.03<br>(0.55, 1.91) | 0.935 | 1.74<br>(0.69, 4.38) | 0.238 | 1.26<br>(0.62, 2.57) | 0.519 |
| 46 – 55 | **1.92**<br>**(1.13, 3.25)** | **0.016** | 1.85<br>(0.99, 3.44) | 0.053 | **2.64**<br>**(1.04, 6.71)** | **0.042** | 0.84<br>(0.39, 1.84) | 0.668 |
| 56 – 65 | **2.74**<br>**(1.61, 4.66)** | **<0.001** | 1.53<br>(0.80, 2.90) | 0.196 | 1.42<br>(0.50, 4.01) | 0.513 | 1.05<br>(0.49, 2.28) | 0.897 |
| >65 | **2.60**<br>**(1.42, 4.76)** | **0.002** | 1.73<br>(0.81, 3.70) | 0.155 | **3.30**<br>**(1.11, 9.83)** | **0.032** | 1.44<br>(0.64, 3.25) | 0.378 |
| Place of residence |  |  |  |  |  |  |  |  |
| Urban | Reference |  |  |  |  |  |  |  |
| Rural | **1.44**<br>**(1.02, 2.02)** | **0.036** | 1.73<br>(0.98, 3.03) | 0.058 | 1.08<br>(0.61, 1.91) | 0.803 | 0.69<br>(0.42, 1.11) | 0.124 |
| Peri-forested | 0.68<br>(0.41, 1.14) | 0.142 | 1.26<br>(0.63, 2.52) | 0.517 | **0.28**<br>**(0.09, 0.86)** | **0.027** | **0.17**<br>**(0.04, 0.33)** | **<0.001** |
| Education in years |  |  |  |  |  |  |  |  |
| >8 years | Reference |  |  |  |  |  |  |  |
| 0 years | 1.12<br>(0.74, 1.69) | 0.606 | 1.35<br>(0.80, 2.27) | 0.262 | 0.58<br>(0.29, 1.15) | 0.120 | 1.53<br>(0.85, 2.76) | 0.159 |
| 1 – 8 years | **1.42**<br>**(1.01, 1.99)** | **0.044** | 1.29<br>(0.80, 2.08) | 0.321 | 1.03<br>(0.60, 1.79) | 0.907 | 1.43<br>(0.86, 2.36) | 0.169 |
| Elevation in meters |  |  |  |  |  |  |  |  |
| >1000 | Reference |  |  |  |  |  |  |  |
| 0 – 500 | **14.16**<br>**(6.43, 31.20)** | **<0.001** | 1.06<br>(0.59, 1.89) | 0.853 | **2.35**<br>**(1.04, 5.31)** | **0.040** | 1.23<br>(0.64, 2.37) | 0.533 |
| 501 – 1000 | **8.39**<br>**(3.72, 18.96)** | **<0.001** | **4.74**<br>**(2.75, 8.17)** | **<0.001** | 0.69<br>(0.24, 2.04) | 0.507 | 1.89<br>(0.94, 3.78) | 0.074 |
| Occupation |  |  |  |  |  |  |  |  |
| Others | Reference |  |  |  |  |  |  |  |
| Farmers | **2.28**<br>**(1.20, 4.32)** | **0.012** | 0.85<br>(0.30, 2.35) | 0.748 | 1.21<br>(0.35, 4.20) | 0.766 | 0.32<br>(0.04, 2.46) | 0.274 |
| Daily wage | 1.39<br>(0.97, 2.01) | 0.075 | 1.60<br>(0.99, 2.59) | 0.055 | 1.28<br>(0.68, 2.41) | 0.444 | 1.48<br>(0.90, 2.44) | 0.121 |
| Housewife | 1.13<br>(0.73, 1.76) | 0.582 | 1.18<br>(0.62, 2.21) | 0.618 | 0.85<br>(0.41, 1.77) | 0.655 | 1.46<br>(0.77, 2.76) | 0.246 |
| Sleeping on |  |  |  |  |  |  |  |  |
| Cot | Reference |  |  |  |  |  |  |  |
| Ground | **1.80**<br>**(1.14, 2.82)** | **0.011** | 1.28<br>(0.77, 2.13) | 0.344 | 1.22<br>(0.54, 2.77) | 0.631 | **2.39**<br>**(1.46, 3.93)** | **0.001** |
| Mat/Mattress | **3.46**<br>**(2.37, 5.04)** | **<0.001** | 1.20<br>(0.74, 1.94) | 0.460 | **2.27**<br>**(1.22, 4.24)** | **0.010** | 0.88<br>(0.52, 1.50) | 0.640 |
| Toilet inside the house |  |  |  |  |  |  |  |  |
| Yes | Reference |  |  |  |  |  |  |  |
| No | **1.49**<br>**(1.12, 1.98)** | **0.006** | **1.58**<br>**(1.10, 2.28)** | **0.014** | 1.24<br>(0.76, 2.04) | 0.388 | 0.78<br>(0.48, 1.27) | 0.313 |
| Changes house clothes daily |  |  |  |  |  |  |  |  |
| Yes | Reference |  |  |  |  |  |  |  |
| No | 1.12<br>(0.78, 1.61) | 0.537 | 0.89<br>(0.51, 1.54) | 0.669 | **3.33**<br>**(1.56, 7.12)** | **0.002** | 0.77<br>(0.42, 1.39) | 0.384 |
| Pet animal |  |  |  |  |  |  |  |  |
| No | Reference |  |  |  |  |  |  |  |
| Yes | 1.30<br>(0.98, 1.72) | 0.072 | **1.72**<br>**(1.22, 2.43)** | **0.002** | **2.53**<br>**(1.58, 4.06)** | **<0.001** | **0.53**<br>**(0.31, 0.89)** | **0.016** |
| **Bush near house** |  |  |  |  |  |  |  |  |
| No | Reference |  |  |  |  |  |  |  |
| Yes | 0.82<br>(0.58, 1.17) | 0.279 | 1.38<br>(0.93, 2.06) | 0.114 | 1.42<br>(0.78, 2.60) | 0.252 | 1.35<br>(0.77, 2.38) | 0.298 |
| **Grass near house** |  |  |  |  |  |  |  |  |
| No | Reference |  |  |  |  |  |  |  |
| Yes | **1.68**<br>**(1.22, 2.32)** | **0.002** | 1.21<br>(0.79, 1.86) | 0.376 | 1.30<br>(0.78, 2.16) | 0.309 | 0.83<br>(0.50, 1.37) | 0.459 |

## Discussion

In this study we aimed to find out the sero-prevalence of the rickettsial disease, and the factors associated with the sero-prevalence. The overall sero-prevalence of ST in this study was 14% which is slightly higher than the reported sero-prevalence of ST in Thailand by Gonwong et al 2022 (22). However, a higher sero-prevalence of 31.8% was reported in Vellore by Trowbridge et al 2017 (23), 30.8% by Khan et al in 2016 in North East India (24) and 19% in Myanmar by Elders et al in 2021 (25). In this research, the sero-prevalence of ST is high in the districts like Tiruvannamalai (22.1%), Vellore (19.8%) and Salem (14.1%) and low in other two districts. This might be due to the variation in the geography and the climatic conditions, as the surveyed areas in these districts are at an elevation of 200 – 1000 MSL and the share the same climate. Even though, Erode shares the same climatic conditions, the land area is well irrigated and due to continuous agricultural activity (26) and less of bushes and shrubs, sero-prevalence of ST might be low. It has been observed in Nepal by Acharya et al 2019, regions with higher elevation and low temperatures have low case burden of scrub typhus (27). The sero-prevalence for scrub typhus was found to be low in the surveyed areas of Nilgiris district. This is because of the low temperature (2 – 25□) and high elevation (1000 to 2600 MSL) of surveyed areas in the Nilgiris.

The sero-prevalence of SF in our participants was 9.1%, which is almost similar to the reported sero-prevalence of 10.1% among North Korean refugees in a study done by Um et al in 2021 (28). On the contrary, higher sero-prevalence of 13.8% in North East India was reported by Khan et al in 2016 (24), 26.2% among Indigenous Populations of Colombia by Oakley et al in 2024 (29) and 29.2% in Mongolia by Ganbold et al in 2023. There are few more studies of SF sero-prevalence reporting as low as 3% among marginalized population from South Carolina by Gual-Gonzalez et al in 2023 and among young men in Thailand by Gonwong et al in 2022 (22). Among the surveyed districts, SF was higher in Salem (19.9%) and in Tiruvannamalai district (13.6%) and lesser in Vellore (5.3%), The Nilgiris (5.2%) and nil in Erode district (0%). The high sero-prevalence in Salem is due to the presence of Kalrayan hills which is at an elevation of 650 – 1000 MSL and Tiruvannamalai due to the presence of Jawadi hills which is at an elevation of 500 – 800 MSL. Similar findings have been observed by Kularatne et al 2003 in hilly areas of Sri Lanka (30).

The sero-prevalence of MT in our study was 3.7% which is almost similar to the 4.2% sero-prevalence reported in North East India (24), Colombia (4.3%) (29), Thailand (4.2%) (31) and in Myanmar (5%) (25). The sero-prevalence of Q fever among the study population was 5.7%, which is slightly lower than the reported 6.9% sero-prevalence by Tshokey et al in 2017 in Bhutan.

Among all the rickettsial infections, females had the higher odds of being sero-prevalence for MT when compared to males. However, other similar studies done in Korea (32) and in Bangladesh (33) have not reported any significant association with gender and MT. There is no statistically significant association between gender and ST, SF and QF positivity. In contrast, many studies had reported that female had a higher odds of being sero-positive for ST in Vellore (17), in Nepal (34), Rajasthan (35) and Korea (36). Whereas for SF, males were at high risk when compared to females in study done in South Carolina (37). Age is found to be a strong risk factor for ST as the risk increases with age in our study. This has also been evident in the 15 year analysis recently published from our centre for Vellore district (17). When compared to people residing in urban areas, people from rural areas were found to be at risk for ST. When compared to people living in peri-forested areas, people from urban areas were at higher risk for MT and QF.

When compared to people living at an elevation of >1000 MSL, people residing at <1000 MSL were having a high sero-prevalence of ST. This agrees with the finding of Tshokey in Bhutan (3). In contrast, a study done at Thailand by Chaisiri et al in 2022, reported a higher sero-prevalence amongst people from upland forest areas for ST (38). People living in 501-1000 MSL were having a high sero-prevalence for SF when compared to the people living at an elevation of >1000 MSL and similar results has been obtained in Bhutan (3). For MT, people living in areas up to 500 MSL were at risk and this as reported by Chaisiri et al in Thailand (38).

Farmers were at high risk for ST in this study and this in agreement with findings from other studies reported from South Korea (39), Guangzhou, China (40) and in Darjeeling, India (41). High sero-prevalence was associated with people sleeping on the ground with or without mat/mattress for ST, which is the same as stated in the study done by Tran et al 2021in Central Vietnam (42). People sleeping on ground with mat/mattress for MT and sleeping on ground without mat/mattress were at higher risk for QF. Not having toilet at home was found to be associated with higher likelihood for ST & SF, the former is in agreement with that observed in Gorakhpur (43). Habit of not changing house clothes daily was associated with high sero-prevalence of MT. Having a pet at home increase the risk for SF and MT seroprevalence. This seems logical as the common pets like cats and dogs can be infested with ticks (SF vectors) and Fleas (MT vectors). In contrast, having a pet decrease the risk of QF. This seems feasible as the major route of transmission of Q Fever is aerosols from infected animals and their products and not by ticks (44,45).

Grassland near house was positively associated with high-prevalence of ST which is also same as reported in the case-control study done by Lyu et al in Beijing, China in 2013 (46). The study limitations include recall bias and single-point data collection. The strengths include adequate sample size across various geographical locations with robust data collection including laboratory testing for IgG antibodies detection for the four rickettsial infections.

In conclusion, sero-prevalence varies across geographical regions and prevalence is associated with certain factors. However, these findings need to be prospectively verified by multi-centric large scale cohort studies done over a period of time.

## Data Availability

All data produced in the present study are available upon reasonable request to the authors

## Author contributions

Study concept and design: J.A.J.P., K.G., S.K.P., S.D. Acquisition of data: J.A.J.P, S.K., S.D. Analysis and interpretation of data: S.D., S.K., K.G., S.K.P., J.A.J.P. Drafting of the manuscript: S.D., S.K., Critical revision of the manuscript: S.K.P., K.G., J.A.J.P. Statistical analysis: S.D., and S.K. All authors have read the finished manuscript; approve and take primary responsibility for the final content.

## Competing interests

The authors declare no competing interests.

Correspondence and requests for materials should be addressed to J.A.J.P

## References

1. Salje J, Weitzel T, Newton PN, Varghese GM, Day N. Rickettsial infections: A blind spot in our view of neglected tropical diseases. PLoS Negl Trop Dis. 2021 May 13;15(5):e0009353. doi:10.1371/journal.pntd.0009353 PubMed PMID: 33983936; PubMed Central PMCID: PMC8118261.

2. Walker DH. Rickettsiae. In: Baron S, editor. Medical Microbiology [Internet]. 4th ed. Galveston (TX): University of Texas Medical Branch at Galveston; 1996 [cited 2024 Oct 2]. Available from: http://www.ncbi.nlm.nih.gov/books/NBK7624/ PubMed PMID: 21413251.

3. Tshokey T, Stenos J, Durrheim DN, Eastwood K, Nguyen C, Graves SR. Seroprevalence of rickettsial infections and Q fever in Bhutan. PLoS Negl Trop Dis. 2017 Nov;11(11):e0006107. doi:10.1371/journal.pntd.0006107 PubMed PMID: 29176880; PubMed Central PMCID: PMC5720829.

4. Sahni SK, Narra HP, Sahni A, Walker DH. Recent molecular insights into rickettsial pathogenesis and immunity. Future Microbiol. 2013 Oct;8(10):1265–88. doi:10.2217/fmb.13.102 PubMed PMID: 24059918; PubMed Central PMCID: PMC3923375.

5. Mittal V, Gupta N, Bhattacharya D, Kumar K, Ichhpujani RL, Singh S, et al. Serological evidence of rickettsial infections in Delhi. Indian J Med Res. 2012 Apr;135(4):538–41. PubMed PMID: 22664504; PubMed Central PMCID: PMC3385240.

6. Acestor N, Cooksey R, Newton PN, Ménard D, Guerin PJ, Nakagawa J, et al. Mapping the Aetiology of Non-Malarial Febrile Illness in Southeast Asia through a Systematic Review— Terra Incognita Impairing Treatment Policies. PLoS ONE. 2012 Sep 6;7(9):e44269. doi:10.1371/journal.pone.0044269 PubMed PMID: 22970193; PubMed Central PMCID: PMC3435412.

7. Abhilash KPP, Jeevan JA, Mitra S, Paul N, Murugan TP, Rangaraj A, et al. Acute Undifferentiated Febrile Illness in Patients Presenting to a Tertiary Care Hospital in South India: Clinical Spectrum and Outcome. J Glob Infect Dis. 2016;8(4):147–54. doi:10.4103/0974-777X.192966 PubMed PMID: 27942194; PubMed Central PMCID: PMC5126753.

8. Devasagayam E, Dayanand D, Kundu D, Kamath MS, Kirubakaran R, Varghese GM. The burden of scrub typhus in India: A systematic review. PLoS Negl Trop Dis. 2021 Jul;15(7):e0009619. doi:10.1371/journal.pntd.0009619 PubMed PMID: 34314437; PubMed Central PMCID: PMC8345853.

9. Parai D, Pattnaik M, Kshatri JS, Rout UK, Peter A, Nanda RR, et al. Scrub typhus seroprevalence from an eastern state of India: findings from the state-wide serosurvey. Trans R Soc Trop Med Hyg. 2023 Jan 1;117(1):22–7. doi:10.1093/trstmh/trac075

10. Sengupta M, Anandan S, Daniel D, Prakash JAJ. Scrub Typhus Seroprevalence in Healthy Indian Population. J Clin Diagn Res JCDR. 2015 Oct;9(10):DM01–2. doi:10.7860/JCDR/2015/14708.6623 PubMed PMID: 26557523; PubMed Central PMCID: PMC4625242.

11. van Eekeren LE, de Vries SG, Wagenaar JFP, Spijker R, Grobusch MP, Goorhuis A. Under-diagnosis of rickettsial disease in clinical practice: A systematic review. Travel Med Infect Dis. 2018 Nov 1;26:7–15. doi:10.1016/j.tmaid.2018.02.006

12. Krishnamoorthi S, Goel S, Kaur J, Bisht K, Biswal M. A Review of Rickettsial Diseases Other Than Scrub Typhus in India. Trop Med Infect Dis. 2023 May 16;8(5):280. doi:10.3390/tropicalmed8050280 PubMed PMID: 37235328; PubMed Central PMCID: PMC10222352.

13. Maurin M, Raoult D. Q Fever. Clin Microbiol Rev. 1999 Oct;12(4):518–53. PubMed PMID: 10515901; PubMed Central PMCID: PMC88923.

14. George T, Rajan SJ, Peter JV, Hansdak SG, Prakash JAJ, Iyyadurai R, et al. Risk Factors for Acquiring Scrub Typhus among the Adults. J Glob Infect Dis. 2018;10(3):147–51. doi:10.4103/jgid.jgid_63_17 PubMed PMID: 30166814; PubMed Central PMCID: PMC6100342.

15. Rose W, Kang G, Verghese VP, Candassamy S, Samuel P, Prakash JJA, et al. Risk factors for acquisition of scrub typhus in children admitted to a tertiary centre and its surrounding districts in South India: a case control study. BMC Infect Dis. 2019 Jul 26;19(1):665. doi:10.1186/s12879-019-4299-2 PubMed PMID: 31349809; PubMed Central PMCID: PMC6660696.

16. Samal C, Das PP, Mishra DK, Sahu DK, Kar PK. Risk Factors Associated with Adult Scrub Typhus Cases in a Tertiary Care Hospital – A Case Control Study. Indian J Community Med. 10.4103/ijcm.ijcm_897_24. doi:10.4103/ijcm.ijcm_897_24

17. D’Cruz S, Sreedevi K, Lynette C, Gunasekaran K, Prakash JAJ. Climate influences scrub typhus occurrence in Vellore, Tamil Nadu, India: analysis of a 15-year dataset. Sci Rep. 2024 Jan 17;14(1):1532. doi:10.1038/s41598-023-49333-5

18. Devamani CS, Schmidt WP, Ariyoshi K, Anitha A, Kalaimani S, Prakash JAJ. Risk Factors for Scrub Typhus, Murine Typhus, and Spotted Fever Seropositivity in Urban Areas, Rural Plains, and Peri-Forest Hill Villages in South India: A Cross-Sectional Study. Am J Trop Med Hyg. 2020 Jul;103(1):238–48. doi:10.4269/ajtmh.19-0642 PubMed PMID: 32458785; PubMed Central PMCID: PMC7356468.

19. Moorthy GS, Rubach MP, Maze MJ, Refuerzo RP, Shirima GM, Lukambagire AS, et al. Prevalence and risk factors for Q fever, spotted fever group rickettsioses, and typhus group rickettsioses in a pastoralist community of northern Tanzania, 2016-2017. Trop Med Int Health TM IH. 2024 May;29(5):365–76. doi:10.1111/tmi.13980 PubMed PMID: 38480005; PubMed Central PMCID: PMC11073910.

20. Vallée J, Thaojaikong T, Moore CE, Phetsouvanh R, Richards AL, Souris M, et al. Contrasting Spatial Distribution and Risk Factors for Past Infection with Scrub Typhus and Murine Typhus in Vientiane City, Lao PDR. PLoS Negl Trop Dis. 2010 Dec 7;4(12):e909. doi:10.1371/journal.pntd.0000909 PubMed PMID: 21151880; PubMed Central PMCID: PMC2998433.

21. D’Cruz S, Perumalla SK, Yuvaraj J, Prakash JAJ. Geography and prevalence of rickettsial infections in Northern Tamil Nadu, India: a cross-sectional study. Sci Rep. 2022 Dec 2;12(1):20798. doi:10.1038/s41598-022-21191-7 PubMed PMID: 36460687; PubMed Central PMCID: PMC9718799.

22. Gonwong S, Mason CJ, Chuenchitra T, Khantapura P, Islam D, Ruamsap N, et al. Nationwide Seroprevalence of Scrub Typhus, Typhus, and Spotted Fever in Young Thai Men. Am J Trop Med Hyg. 2022 Apr 4;106(5):1363–9. doi:10.4269/ajtmh.20-1512 PubMed PMID: 35378507; PubMed Central PMCID: PMC9128670.

23. Trowbridge P P. D, Premkumar PS, Varghese GM. Prevalence and risk factors for scrub typhus in South India. Trop Med Int Health. 2017;22(5):576–82. doi:10.1111/tmi.12853

24. Khan SA, Bora T, Chattopadhyay S, Jiang J, Richards AL, Dutta P. Seroepidemiology of rickettsial infections in Northeast India. Trans R Soc Trop Med Hyg. 2016 Aug 1;110(8):487–94. doi:10.1093/trstmh/trw052

25. Elders PND, Swe MMM, Phyo AP, McLean ARD, Lin HN, Soe K, et al. Serological evidence indicates widespread distribution of rickettsioses in Myanmar. Int J Infect Dis IJID Off Publ Int Soc Infect Dis. 2021 Feb;103:494–501. doi:10.1016/j.ijid.2020.12.013 PubMed PMID: 33310022; PubMed Central PMCID: PMC7862081.

26. INDUSTRIAL POTENTIAL SURVEY REPORT - ERODE [Internet]. [cited 2024 Aug 22]. Available from: https://dcmsme.gov.in/publications/traderep/erode/erode2.htm

27. Acharya BK, Chen W, Ruan Z, Pant GP, Yang Y, Shah LP, et al. Mapping Environmental Suitability of Scrub Typhus in Nepal Using MaxEnt and Random Forest Models. Int J Environ Res Public Health. 2019 Dec;16(23):4845. doi:10.3390/ijerph16234845 PubMed PMID: 31810239; PubMed Central PMCID: PMC6926588.

28. Um J, Nam Y, Lim JN, Kim M, An Y, Hwang SH, et al. Seroprevalence of scrub typhus, murine typhus and spotted fever groups in North Korean refugees. Int J Infect Dis IJID Off Publ Int Soc Infect Dis. 2021 May;106:23–8. doi:10.1016/j.ijid.2021.02.111 PubMed PMID: 33676004.

29. Oakley R, Kann S, Concha G, Plag M, Poppert S, Graves S, et al. Seroprevalence of Rickettsia Spp. and Orientia tsutsugamushi in Indigenous Populations from the Sierra Nevada de Santa Marta, Colombia. Vector Borne Zoonotic Dis Larchmt N. 2024 May 14. doi:10.1089/vbz.2023.0077 PubMed PMID: 38742967.

30. Kularatne S a. M, Edirisingha JS, Gawarammana IB, Urakami H, Chenchittikul M, Kaiho I. Emerging rickettsial infections in Sri Lanka: the pattern in the hilly Central Province. Trop Med Int Health TM IH. 2003 Sep;8(9):803–11. doi:10.1046/j.1365-3156.2003.01108.x PubMed PMID: 12950666.

31. Bhengsri S, Baggett HC, Edouard S, Dowell SF, Dasch GA, Fisk TL, et al. Sennetsu Neorickettsiosis, Spotted Fever Group, and Typhus Group Rickettsioses in Three Provinces in Thailand. Am J Trop Med Hyg. 2016 Jul 6;95(1):43–9. doi:10.4269/ajtmh.15-0752 PubMed PMID: 27139448; PubMed Central PMCID: PMC4944706.

32. Chang BJ, Kim SJ, Lee WC, Lee MJ, Choe NH. Comparative Study on the Epidemiological Trends and Aspects of Murine Typhus in Korea in the Last Decade (2006-2015). J Glob Infect Dis. 2018;10(3):121–4. doi:10.4103/jgid.jgid_140_17 PubMed PMID: 30166809; PubMed Central PMCID: PMC6100331.

33. Maude RR, Maude RJ, Ghose A, Amin MR, Islam MB, Ali M, et al. Serosurveillance of Orientia tsutsugamushi and Rickettsia typhi in Bangladesh [Internet]. 2014 Sep 3. doi:10.4269/ajtmh.13-0570

34. Gautam R, Parajuli K, Sherchand JB. Epidemiology, Risk Factors and Seasonal Variation of Scrub Typhus Fever in Central Nepal. Trop Med Infect Dis. 2019 Mar;4(1):1. doi:10.3390/tropicalmed4010027

35. Bithu R, Kanodia V, Maheshwari R. Possibility of scrub typhus in fever of unknown origin (FUO) cases: An experience from Rajasthan. Indian J Med Microbiol. 2014 Oct 1;32(4):387–90. doi:10.4103/0255-0857.142241

36. Bang HA, Lee MJ, Lee WC. Comparative Research on Epidemiological Aspects of Tsutsugamushi Disease (Scrub Typhus) between Korea and Japan. Jpn J Infect Dis. 2008;61(2):148–50. doi:10.7883/yoken.JJID.2008.148

37. Gual-Gonzalez L, Self SCW, Meyer M, Cantillo-Barraza O, Torres ME, Nolan MS. Human spotted fever group Rickettsia seroprevalence and associated epidemiologic factors among diverse, marginalized populations in South Carolina. Ticks Tick-Borne Dis. 2024 Mar;15(2):102288. doi:10.1016/j.ttbdis.2023.102288 PubMed PMID: 38071922.

38. Chaisiri K, Tanganuchitcharnchai A, Kritiyakan A, Thinphovong C, Tanita M, Morand S, et al. Risk factors analysis for neglected human rickettsioses in rural communities in Nan province, Thailand: A community-based observational study along a landscape gradient. PLoS Negl Trop Dis. 2022 Mar 23;16(3):e0010256. doi:10.1371/journal.pntd.0010256

39. Park JH, Gill B, Acharya D, Yoo SJ, Lee K, Lee J. Seroprevalence and Factors Associated with Scrub Typhus Infection among Forestry Workers in National Park Offices in South Korea. Int J Environ Res Public Health. 2021 Mar 18;18(6):3131. doi:10.3390/ijerph18063131 PubMed PMID: 33803616; PubMed Central PMCID: PMC8003109.

40. Lu J, Liu Y, Ma X, Li M, Yang Z. Impact of Meteorological Factors and Southern Oscillation Index on Scrub Typhus Incidence in Guangzhou, Southern China, 2006–2018. Front Med. 2021 Jul 28;8:667549. doi:10.3389/fmed.2021.667549 PubMed PMID: 34395468; PubMed Central PMCID: PMC8355740.

41. Sharma PK, Ramakrishnan R, Hutin YJF, Barui AK, Manickam P, Kakkar M, et al. Scrub typhus in Darjeeling, India: opportunities for simple, practical prevention measures⍰. Trans R Soc Trop Med Hyg. 2009 Nov 1;103(11):1153–8. doi:10.1016/j.trstmh.2009.02.006

42. Tran HTD, Hattendorf J, Do HM, Hoang TT, Hoang HTH, Lam HN, et al. Ecological and behavioural risk factors of scrub typhus in central Vietnam: a case-control study. Infect Dis Poverty. 2021 Aug 19;10:110. doi:10.1186/s40249-021-00893-6 PubMed PMID: 34412700; PubMed Central PMCID: PMC8374119.

43. Thangaraj JWV, Vasanthapuram R, Machado L, Arunkumar G, Sodha SV, Zaman K, et al. Risk Factors for Acquiring Scrub Typhus among Children in Deoria and Gorakhpur Districts, Uttar Pradesh, India, 2017. Emerg Infect Dis. 2018 Dec;24(12):2364–7. doi:10.3201/eid2412.180695 PubMed PMID: 30457537; PubMed Central PMCID: PMC6256400.

44. Schimmer B, Schotten N, van Engelen E, Hautvast JLA, Schneeberger PM, van Duijnhoven YTHP. Coxiella burnetii Seroprevalence and Risk for Humans on Dairy Cattle Farms, the Netherlands, 2010–2011. Emerg Infect Dis. 2014 Mar;20(3):417–25. doi:10.3201/eid2003.131111 PubMed PMID: 24572637; PubMed Central PMCID: PMC3944848.

45. Duron O, Sidi-Boumedine K, Rousset E, Moutailler S, Jourdain E. The Importance of Ticks in Q Fever Transmission: What Has (and Has Not) Been Demonstrated? Trends Parasitol. 2015 Nov 1;31(11):536–52. doi:10.1016/j.pt.2015.06.014

46. Lyu Y, Tian L, Zhang L, Dou X, Wang X, Li W, et al. A Case-Control Study of Risk Factors Associated with Scrub Typhus Infection in Beijing, China. PLOS ONE. 2013 May 14;8(5):e63668. doi:10.1371/journal.pone.0063668

