## Supplementary file for "Factors associated with the sero-prevalence of Rickettsioses in Northern Tamil Nadu, India"

### Supplementary data

**Supplementary Table 1: Prevalence of ST, SF, MT and Q fever**

| District | Total tested | ST | | SF | | MT | | Q fever | |
|---|---|---|---|---|---|---|---|---|---|
| | | n | % | n | % | n | % | n | % |
| Erode | 406 | 9 | 2.2% | 0 | 0% | 9 | 2.2% | 26 | 6.4% |
| The Nilgiris | 345 | 7 | 2.0% | 18 | 5.2% | 7 | 2% | 15 | 4.3% |
| Salem | 512 | 72 | 14.1% | 102 | 19.9% | 9 | 1.8% | 59 | 11.5% |
| Tiruvannamalai | 535 | 118 | 22.1% | 73 | 13.6% | 56 | 10.5% | 7 | 1.3% |
| Vellore | 767 | 152 | 19.8% | 41 | 5.3% | 13 | 1.7% | 39 | 5.1% |
| **Total** | **2565** | **358** | **14%** | **234** | **9.1%** | **94** | **3.7%** | **146** | **5.7%** |

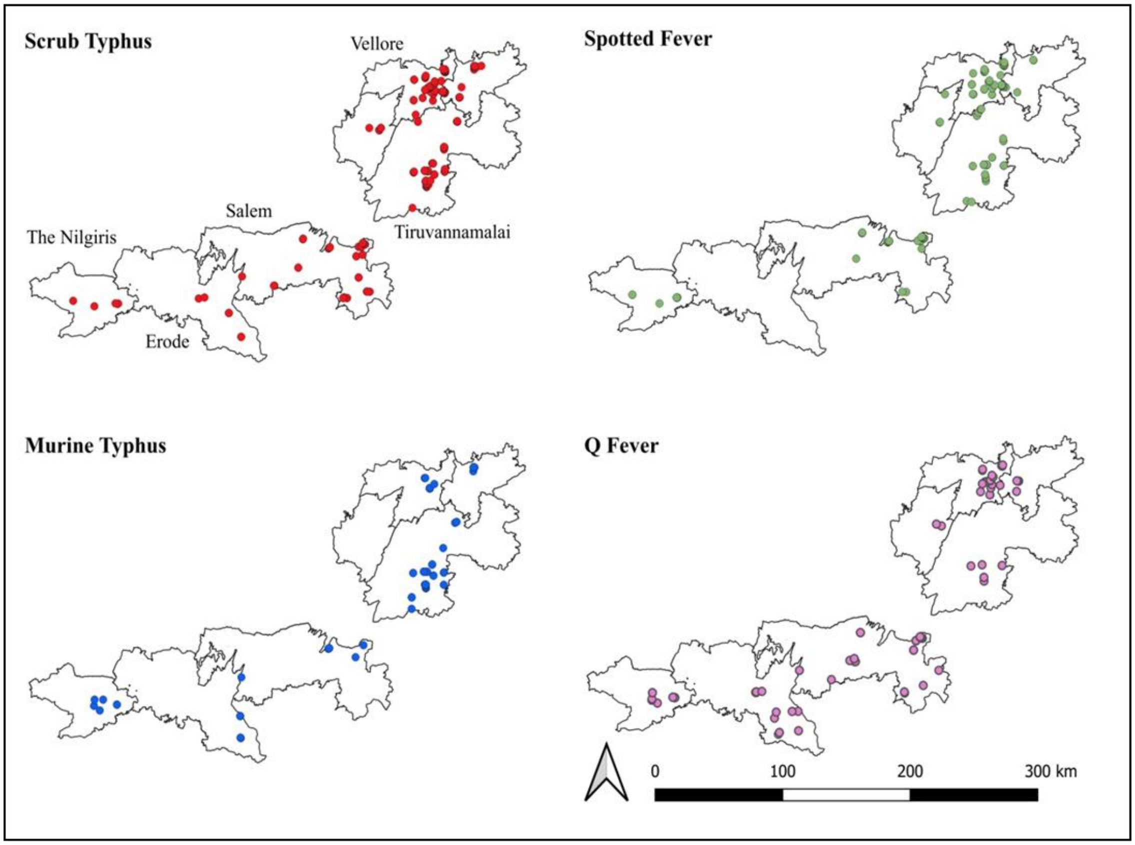


**Supplementary Figure 1:** Map of the surveyed study districts with the rickettsia positives geo-coded

The maximum number of scrub typhus IgG positives are from Vellore, Tiruvannamalai and least in The Nilgiris and Erode districts. Whereas for SF, Salem and Tiruvannamalai reported more cases and Erode reported zero SF. Tiruvannamalai has reported a high number of MT cases and The Nilgiris with a minimum number of cases. Q fever numbers were highest in Salem and lowest in Tiruvannamalai.

**Supplementary Table 2: Demographics characteristics of participants**

| Variable (n=2565) | Category | Frequency | Percentage (%) |
|---|---|---|---|
| **Sex** | Male | 1025 | 40 |
| | Female | 1540 | 60 |
| **Age** | <=30 | 641 | 25 |
| | 31-45 | 807 | 31.5 |
| | 46-60 | 692 | 27 |
| | >60 | 425 | 16.5 |
| **Occupation** | Daily wage workers | 1286 | 50.1 |
| | Farmer | 95 | 3.7 |
| | House wife | 539 | 21 |
| | Students | 170 | 6.6 |
| | Unemployed | 245 | 9.6 |
| | Other | 230 | 9 |
| **Education** | Illiterate | 884 | 34.5 |
| | Primary | 384 | 15 |
| | Middle | 438 | 17.1 |
| | High | 409 | 15.9 |
| | +2 | 200 | 7.8 |
| | Graduate | 234 | 9.1 |
| | PG | 16 | 0.6 |
| **Locality** | Urban | 849 | 33.1 |
| | Rural | 1302 | 50.8 |
| | Peri-forested | 414 | 16.1 |
| **Elevation** | 0 – 500m | 1723 | 67.2 |
| | 501 – 1000m | 495 | 19.3 |
| | >1000m | 347 | 13.5 |
| **Districts surveyed** | Erode | 406 | 15.8 |
| | The Nilgiris | 345 | 13.5 |
| | Salem | 512 | 19.9 |
| | Tiruvannamalai | 535 | 20.9 |
| | Vellore | 767 | 29.9 |

**Supplementary Table 3: Factors associated with the prevalence of ST**

| Variables (n = 2565) | | Scrub typhus | | OR | 95% CI | p Value |
|---|---|---|---|---|---|---|
| | | **Positive** | **Negative** | | | |
| **Gender** | Female | 234 (15.2%) | 1306 (84.8%) | **1.30** | **1.03, 1.65** | **0.027** |
| | Male | 124 (12.1%) | 901 (87.9%) | | | |
| **Age group in years** | 16 –25 | 29 (80%) | 335 (92.5%) | Reference | | |
| | 26 – 35 | 46 (8.8%) | 474 (91.2%) | 1.12 | 0.69, 1.82 | 0.644 |
| | 36 – 45 | 88 (15.6%) | 476 (84.4%) | **2.14** | **1.37, 3.32** | **0.001** |
| | 46 – 55 | 77 (16.8%) | 382 (83.2%) | **2.33** | **1.48, 3.66** | **<0.001** |
| | 56 – 65 | 83 (19.2%) | 349 (80.8%) | **2.75** | **1.75, 4.30** | **<0.001** |
| | >65 | 35 (15.5%) | 191 (84.5%) | **2.12** | **1.25, 3.57** | **0.005** |
| **Place of residence** | Urban | 87 (10.2%) | 762 (89.8%) | Reference | | |
| | Rural | 218 (16.7%) | 1084 (83.3%) | **1.76** | **1.35, 2.30** | **<0.001** |
| | Peri-forested | 53 (12.8%) | 361 (87.2%) | 1.29 | 0.89, 1.85 | 0.175 |
| **Education in years** | >8 | 79 (9.2%) | 780 (90.8%) | Reference | | |
| | 0 | 126 (14.3%) | 758 (85.7%) | **1.64** | **1.22, 2.21** | **0.001** |
| | 1 – 8 | 153 (18.6%) | 669 (81.4%) | **2.26** | **1.69, 3.02** | **<0.001** |
| **Elevation in meters** | >1000 | 7 (2%) | 340 (98%) | Reference | | |
| | 0 – 500 | 280 (16.3%) | 1443 (83.7%) | **9.43** | **4.41, 20.14** | **<0.001** |
| | 501 – 1000 | 71 (14.3%) | 424 (85.7%) | **8.13** | **3.69, 17.91** | **<0.001** |
| **Occupation** | Others | 60 (9.3%) | 585 (90.7%) | Reference | | |
| | Farmers | 21 (22.1%) | 74 (77.9%) | **2.77** | **1.59, 4.81** | **<0.001** |
| | Daily wage | 195 (15.2%) | 1091 (84.8%) | **1.74** | **1.28, 2.37** | **<0.001** |
| | House wife | 82 (15.2%) | 457 (84.8%) | **1.75** | **1.23, 2.50** | **<0.001** |
| **Sleeping on** | Cot | 46 (6.4%) | 668 (93.6%) | Reference | | |
| | Ground | 63 (10.5%) | 537 (89.5%) | **1.70** | **1.15, 2.53** | **0.008** |
| | Mat/Mattress | 249 (19.9%) | 1002 (80.1%) | **3.61** | **2.60, 5.02** | **<0.001** |
| **Toilet inside the house** | No | 169 (18%) | 769 (82%) | **1.67** | **1.34, 2.09** | **<0.001** |
| | Yes | 189 (11.6%) | 1438 (88.4%) | | | |
| **Changes house clothes daily** | No | 271 (12.8%) | 1852 (87.2%) | **0.60** | **0.46, 0.78** | **<0.001** |
| | Yes | 87 (19.7%) | 355 (80.3%) | | | |
| **Pet animal** | Yes | 117 (18.7%) | 509 (81.3%) | **1.62** | **1.27, 2.06** | **<0.001** |
| | No | 241 (12.4%) | 1698 (87.6%) | | | |
| **Bush near house** | Yes | 94 (19%) | 401 (81%) | **1.60** | **1.24, 2.08** | **<0.001** |
| | No | 264 (12.8%) | 1806 (87.2%) | | | |
| **Grass near house** | Yes | 194 (16.3%) | 995 (83.7%) | **1.44** | **1.15, 1.80** | **0.001** |
| | No | 164 (11.9%) | 1212 (88.1%) | | | |

**Supplementary Table 4: Factors associated with the prevalence of SF**

| **Variables (n = 2565)** | | **Spotted fever** | | **OR** | **95% CI** | **p Value** |
|---|---|---|---|---|---|---|
| | | **Positive** | **Negative** | | | |
| **Gender** | Female | 133 (8.6%) | 1407 (91.4%) | 0.87 | 0.66, 1.14 | 0.327 |
| | Male | 101 (9.9%) | 924 (90.1%) | | | |
| **Age group in years** | 16 –25 | 20 (5.5%) | 344 (94.5%) | Reference | | |
| | 26 – 35 | 50 (9.6%) | 470 (90.4%) | 1.83 | 1.07, 3.13 | 0.027 |
| | 36 – 45 | 43 (7.6%) | 521 (92.4%) | 1.42 | 0.82, 2.46 | 0.210 |
| | 46 – 55 | 56 (12.2%) | 403 (87.8%) | **2.39** | **1.41, 4.06** | **0.001** |
| | 56 – 65 | 47 (10.9%) | 385 (89.1%) | **2.10** | **1.22, 3.61** | **0.007** |
| | >65 | 18 (8%) | 208 (92%) | 1.49 | 0.77, 2.88 | 0.237 |
| **Place of residence** | Urban | 21 (2.5%) | 828 (97.5%) | Reference | | |
| | Rural | 158 (12.1%) | 1144 (87.9%) | 5.45 | **3.42, 8.67** | **<0.001** |
| | Peri-forested | 55 (13.3%) | 359 (86.7%) | 6.04 | **3.60, 10.14** | **<0.001** |
| **Education in years** | >8 | 36 (4.2%) | 823 (95.8%) | Reference | | |
| | 0 | 133 (15%) | 751 (85%) | **4.05** | **2.77, 5.93** | **<0.001** |
| | 1 – 8 | 65 (7.9%) | 757 (92.1%) | **1.96** | **1.29, 2.99** | **0.002** |
| **Elevation in meters** | >1000 | 19 (5.5%) | 328 (94.5%) | Reference | | |
| | 0 – 500 | 76 (4.4%) | 1647 (95.6%) | 0.80 | 0.48, 1.34 | 0.388 |
| | 501 – 1000 | 139 (28.1%) | 356 (71.9%) | **6.74** | **4.08, 11.14** | **<0.001** |
| **Occupation** | Others | 28 (4.3%) | 617 (95.7%) | Reference | | |
| | Farmers | 5 (5.3%) | 90 (94.7%) | 1.22 | 0.46, 3.25 | 0.685 |
| | Daily wage | 174 (13.5%) | 1112 (86.5%) | **3.45** | **2.29, 5.20** | **<0.001** |
| | House wife | 27 (5%) | 512 (95%) | 1.16 | 0.68, 2.00 | 0.587 |
| **Sleeping on** | Cot | 31 (4.3%) | 683 (95.7%) | Reference | | |
| | Ground | 99 (16.5%) | 501 (83.5%) | **4.35** | **2.86, 6.62** | **<0.001** |
| | Mat/Mattress | 104 (8.3%) | 1147 (91.7%) | **2.00** | **1.32, 3.02** | **0.001** |
| **Toilet inside the house** | No | 152 (16.2%) | 786 (83.8%) | **3.64** | **2.75, 4.83** | **<0.001** |
| | Yes | 82 (5%) | 1545 (95%) | | | |
| **Changes house clothes daily** | No | 210 (9.9%) | 1913 (90.1%) | **1.91** | **1.24, 2.96** | **0.004** |
| | Yes | 24 (5.4%) | 418 (94.6%) | | | |
| **Pet animal** | Yes | 72 (11.5%) | 554 (88.5%) | **1.43** | **1.06, 1.91** | **0.020** |
| | No | 162 (8.4%) | 1777 (91.6%) | | | |
| **Bush near house** | Yes | 80 (16.2%) | 415 (83.8%) | **2.40** | **1.79, 3.21** | **<0.001** |
| | No | 154 (7.4%) | 1916 (92.6%) | | | |
| **Grass near house** | Yes | 162 (13.6%) | 1027 (86.4%) | **2.86** | **2.14, 3.82** | **<0.001** |
| | No | 72 (5.2%) | 1304 (94.8%) | | | |

**Supplementary Table 5: Factors associated with the prevalence of MT**

| Variables (n = 2565) | | Murine typhus | | OR | 95% CI | p Value |
|---|---|---|---|---|---|---|
| | | Positive | Negative | | | |
| **Gender** | Female | 72 (4.7%) | 1468 (95.3%) | **2.24** | **1.38, 3.63** | **0.001** |
| | Male | 22 (2.1%) | 1003 (97.9%) | | | |
| **Age group in years** | 16 –25 | 8 (2.2%) | 356 (97.8%) | Reference | | |
| | 26 – 35 | 22 (4.2%) | 498 (95.8%) | 1.97 | 0.87, 4.47 | 0.106 |
| | 36 – 45 | 21 (3.7%) | 543 (96.3%) | 1.72 | 0.75, 3.93 | 0.197 |
| | 46 – 55 | 23 (5%) | 436 (95%) | **2.35** | **1.04, 5.31** | **0.041** |
| | 56 – 65 | 11 (2.5%) | 421 (97.5%) | 1.16 | 0.46, 2.92 | 0.749 |
| | >65 | 9 (4%) | 217 (96%) | 1.85 | 0.70, 4.86 | 0.214 |
| **Place of residence** | Urban | 30 (3.5%) | 819 (96.5%) | Reference | | |
| | Rural | 59 (4.5%) | 1243 (95.5%) | 1.30 | 0.83, 2.03 | 0.257 |
| | Peri-forested | 5 (1.2%) | 409 (98.8%) | **0.33** | **0.13, 0.87** | **0.024** |
| **Education in years** | >8 | 32 (3.7%) | 827 (96.3%) | Reference | | |
| | 0 | 24 (2.7%) | 860 (97.3%) | 0.72 | 0.42, 1.24 | 0.234 |
| | 1 – 8 | 38 (4.6%) | 784 (95.4%) | 1.25 | 0.78, 2.03 | 0.358 |
| **Elevation in meters** | >1000 | 8 (2.3%) | 339 (97.7%) | Reference | | |
| | 0 – 500 | 79 (4.6%) | 1644 (95.4%) | 2.04 | 0.98, 4.25 | 0.058 |
| | 501 – 1000 | 7 (1.4%) | 488 (98.6%) | 0.61 | 0.22, 1.69 | 0.341 |
| **Occupation** | Others | 19 (2.9%) | 626 (97.1%) | Reference | | |
| | Farmers | 4 (4.2%) | 91 (95.8%) | 1.45 | 0.48, 4.35 | 0.509 |
| | Daily wage | 48 (3.7%) | 1238 (96.3%) | 1.28 | 0.75, 2.19 | 0.374 |
| | House wife | 23 (4.3%) | 516 (95.7%) | 1.47 | 0.79, 2.73 | 0.223 |
| **Sleeping on** | Cot | 16 (2.2%) | 698 (97.8%) | Reference | | |
| | Ground | 12 (2%) | 588 (98%) | 0.89 | 0.42, 1.90 | 0.763 |
| | Mat/Mattress | 66 (5.3%) | 1185 (94.7%) | **2.43** | **1.40, 4.23** | **0.002** |
| **Toilet inside the house** | No | 36 (3.8%) | 902 (96.2%) | 1.08 | 0.71, 1.65 | 0.744 |
| | Yes | 58 (3.6%) | 1569 (96.4%) | | | |
| **Changes house clothes daily** | No | 83 (3.9%) | 2040 (96.1%) | 1.59 | 0.84, 3.02 | 0.165 |
| | Yes | 11 (2.5%) | 431 (97.5%) | | | |
| **Pet animal** | Yes | 50 (8%) | 576 (92%) | **3.74** | **2.47, 5.67** | **<0.001** |
| | No | 44 (2.3%) | 1895 (97.7%) | | | |
| **Bush near house** | Yes | 23 (4.6%) | 472 (95.4%) | 1.37 | 0.85, 2.22 | 0.230 |
| | No | 71 (3.4%) | 1999 (96.6%) | | | |
| **Grass near house** | Yes | 43 (3.6%) | 1146 (96.4%) | 0.98 | 0.65, 1.47 | 0.917 |
| | No | 51 (3.7%) | 1325 (96.3%) | | | |

**Supplementary Table 6: Factors associated with the prevalence of QF**

| Variables (n = 2565) | | Q Fever | | OR | 95% CI | p Value |
|---|---|---|---|---|---|---|
| | | Positive | Negative | | | |
| **Gender** | Female | 81 (5.3%) | 1459 (94.7%) | 0.82 | 0.59, 1.15 | 0.259 |
| | Male | 65 (6.3%) | 960 (93.7%) | | | |
| **Age group in years** | 16 –25 | 14 (3.8%) | 350 (96.2%) | Reference | | |
| | 26 – 35 | 24 (4.6%) | 496 (95.4%) | 1.21 | 0.62, 2.37 | 0.579 |
| | 36 – 45 | 39 (6.9%) | 525 (93.1%) | 1.86 | 0.99, 3.47 | 0.052 |
| | 46 – 55 | 23 (5%) | 436 (95%) | 1.32 | 0.67, 2.60 | 0.424 |
| | 56 – 65 | 27 (6.3%) | 405 (93.8%) | 1.67 | 0.86, 3.23 | 0.130 |
| | >65 | 19 (8.4%) | 207 (91.6%) | **2.30** | **1.13, 4.67** | **0.022** |
| **Place of residence** | Urban | 61 (7.2%) | 788 (92.8%) | Reference | | |
| | Rural | 80 (6.1%) | 1222 (93.9%) | 0.85 | 0.60, 1.19 | 0.341 |
| | Peri-forested | 5 (1.2%) | 409 (98.8%) | **0.16** | **0.06, 0.40** | **<0.001** |
| **Education in years** | >8 | 35 (4.1%) | 824 (95.9%) | Reference | | |
| | 0 | 61 (6.9%) | 823 (93.1%) | **1.75** | **1.14, 2.67** | **0.011** |
| | 1 – 8 | 50 (6.1%) | 772 (93.9%) | 1.53 | 0.98, 2.38 | 0.062 |
| **Elevation in meters** | >1000 | 16 (4.6%) | 331 (95.4%) | Reference | | |
| | 0 – 500 | 92 (5.3%) | 1631 (94.7%) | 1.17 | 0.68, 2.01 | 0.578 |
| | 501 – 1000 | 38 (7.7%) | 457 (92.3%) | 1.72 | 0.94, 3.14 | 0.077 |
| **Occupation** | Others | 31 (4.8%) | 614 (95.2%) | Reference | | |
| | Farmers | 1 (1.1%) | 94 (98.9%) | 0.21 | 0.03, 1.56 | 0.128 |
| | Daily wage | 87 (6.8%) | 1199 (93.2%) | 1.44 | 0.94, 2.19 | 0.092 |
| | House wife | 27 (5%) | 512 (95%) | 1.04 | 0.62, 1.77 | 0.872 |
| **Sleeping on** | Cot | 36 (5%) | 678 (95%) | Reference | | |
| | Ground | 59 (9.8%) | 541 (90.2%) | **2.05** | **1.34, 3.16** | **0.001** |
| | Mat/Mattress | 51 (4.1%) | 1200 (95.9%) | 0.80 | 0.52, 1.24 | 0.318 |
| **Toilet inside the house** | No | 45 (4.8%) | 893 (95.2%) | 0.76 | 0.53, 1.09 | 0.157 |
| | Yes | 101 (6.2%) | 1526 (93.8%) | | | |
| **Changes house clothes daily** | No | 118 (5.6%) | 2005 (94.4%) | 0.87 | 0.57, 1.33 | 0.572 |
| | Yes | 28 (6.3%) | 414 (93.7%) | | | |
| **Pet animal** | Yes | 19 (3%) | 607 (97%) | **0.45** | **0.27, 0.73** | **0.001** |
| | No | 127 (6.5%) | 1812 (93.5%) | | | |
| **Bush near house** | Yes | 36 (7.3%) | 459 (92.7%) | 1.40 | 0.95, 2.06 | 0.105 |
| | No | 110 (5.3%) | 1960 (94.7%) | | | |
| **Grass near house** | Yes | 60 (5%) | 1129 (95%) | 0.80 | 0.57, 1.12 | 0.797 |
| | No | 86 (6.3%) | 1290 (93.8%) | | | |
